# Reopening Italy’s Schools in September 2020: A Bayesian Estimation of the Change in the Growth Rate of New SARS-CoV-2 Cases

**DOI:** 10.1101/2021.04.06.21254993

**Authors:** Luca Casini, Marco Roccetti

## Abstract

**Objectives:** CoViD-19’s second wave started a debate on the potential role of schools as a primary factor in the contagion resurgence. Two opposite positions appeared: those convinced that schools played a major role in spreading SARS-CoV-2 infections and those who were not. We studied the growth rate of the total number of SARS-CoV-2 infections in all the Italian regions, before and after the school reopening (September - October 2020), investigating the hypothesis of an association between schools and the resurgence of the virus in Italy.

**Methods:** Using Bayesian piecewise linear regression to scrutinize the number of daily SARS-CoV-2 infections in each Italian, we looked for an estimate of a changepoint in the growth rate of those confirmed cases. We compared the changepoints with the school opening dates, for each Italian region. The regression allows to discuss the change in steepness of the infection curve, before and after the changepoint.

**Results:** In 15 out of 21 Italian regions (71%), an estimated change in the rate of growth of the total number of daily SARS-CoV-2 infection cases occurred after an average of 16.66 days (CI 95% 14.47 to 18.73) since the school reopening. The number of days required for the SARS-CoV-2 daily cases to double went from an average of 47.50 days (CI 95% 37.18 to 57.61) before the changepoint to an average of 7.72 days (CI 95% 7.00 to 8.48) after it.

**Conclusion:** Studying the rate of growth of daily SARS-CoV-2 cases in all the Italian regions provides some evidence in favor of a link between school reopening and the resurgence of the virus in Italy. The number of factors that could have played a role are too many to give a definitive answer. Still, the temporal correspondence warrants for a controlled experiment to clarify how much reopening schools mattered.

**Design:** Not Applicable

**Setting:** Not Applicable

**Participants:** Not Applicable

**Interventions:** Not Applicable

**Primary and Secondary Outcome Measures:** Not Applicable

**Article Summary:** *Strengths and Limitations of this Study:*

- The use of a Bayesian linear regression model represents a reliable method to account for the uncertainty in the estimation of the changes of the growth rate of the number of daily SARS-CoV-2 infections.
- Analyzing the variation of the total number of new daily SARS-CoV-2 confirmed cases per each Italian region, in coincidence with school reopening, has avoided the problems of looking for specific data collected in schools.
- The problem has been avoided of many infections missed inside schools as positive children and adolescents tend to display less symptoms, therefore leading to a lower probability to be detected with a passive surveillance methodology.
- Data, made available by the Italian Government, used to count the number of daily SARS-CoV-2 infections, amount to aggregated measures, uploaded daily: in some cases, those measures changed meaning over time and/or contained errors that were never corrected.
- Many confounding factors, besides schools, may have played a role, nonetheless in September 2020 in Italy those factors were still fewer than in the following months, when several various containment measures were put in place.

**Author Contribution:** LC and MR equally contributed to conceive, design, write, manage, and revise the manuscript.

**Funding:** The authors have not declared a specific grant for this research from any funding agency in the public, commercial or not-for-profit sectors.

**Competing interests:** None declared.

**Patient consent for publication:** Not required.

**Provenance and peer review:** Not commissioned, externally peer reviewed.

**Data availability statement:** Data are available in a public, open access repository (https://github.com/pcm-dpc/COVID-19). All data and code are available upon request to the corresponding author

**Research Ethics Approval: Human Participants:** Not applicable: neither humans nor animals nor personal data are being involved in this study.

## Introduction

We recognize that attending schools play a key role for younger generations in supporting their development toward shared societal values and in promoting their positive physical and mental wellbeing. Nonetheless, if the scientific community is to overcome the challenges posed by SARS-CoV-2, a serious assessment of the impact that educational settings may have had in the spread of the pandemic cannot take a back seat. Hence the main motivation of our study.

As the summer of 2020 was coming to its end, with its relatively low number of CoViD-19 cases in most western countries, many started arguing whether it was wise to normally restart school activities. During the first wave, in most nations schools were closed, as any other activity, and only some partially reopened them as the situation got under control and the lockdown was lifted. In this context, the large CoViD-19 outbreak in a high school in Israel in May 2020 ignited the discussions about the role of schools in the spread of the virus [1,2]. Two opposing sides appeared quite clearly: on one side, those who consider schools to be a minor risk and the importance of school paramount; on the other, we have those who are concerned by the lack of clear data on the contagion dynamics in schools and are scared by the high number of asymptomatic cases in younger people. The same discussion obviously emerged in all the other countries affected by the ongoing pandemic, with the two sides bringing to the table mostly the same arguments with the occasional country specific remarks.

Most international literature that suggests the absence of considerable risk factors connected to schools focuses on the fact that children and adolescents seem to be the least affected by the virus, both in terms of the number of positive cases but also of symptoms and contagiousness. Ismail et al., studying UK schools, show how the incidence in students is correlated to the total incidence in the region and how the most cases inside schools are transmission between staff members [3]. Similar studies targeting other countries draw similar conclusions, especially when it comes to primary schools and kindergarten [4–9]. The hypothesis that children could be super-spreaders was put forward, as a large asymptomatic population could easily and unknowingly spread the virus in their families. Munro and Faust, however highlight how there is no evidence supporting this idea [10]. Viner et al. in their survey find that children have a lower susceptibility to the virus and thus play a lesser role in the transmission of it[11]. Similar results emerge from Ludvigsson [12]. Cheng et al. use the Taiwanese example to propose how to reopen universities safely [13].

The researchers presenting the opposite hypothesis are pointing to the weaknesses of many of the aforementioned studies, namely the very small samples and the fact that often the role of asymptomatic subjects is not tracked and considered properly. Many point out the correspondence between the insurgence of the second wave in many countries within two weeks from the school openings and point to the data that suggest a higher spread in the school-aged population in those months, especially in high school and university students [14,15]. Flasche and Edmunds respond to Ismail’s study saying that it was conducted with schools not fully populated and underestimates the potential of children, especially in the 10-18 age bracket [16]. This group has seen a considerable increase in September, as did college students, and seemed to be a common source of SARS-CoV-2 infections in the households. Yamey and Walensky [17] express their concerns for universities reopening. They also note how blaming young adults for their actions is an ineffective public health strategy and advise for a harm-reduction approach, with effort put into effective communication, safe events and guidance specifically tailored for young people. Sebastiani and Palù [18] study the Italian situation and argue that the rise of new cases in September, with most SARS-CoV-2 infections happening in the household, is compatible with the hypothesis of school being a factor. While, inside schools, measures were taken, they argue that outside contact was inevitable due to public transportation and social gatherings, so that young people spread the virus among themselves. Larosa et al. [19], conducted a study in the Reggio Emilia province (Emilia-Romagna region, Italy) showing that there were non-negligible clusters in the age bracket 10-18 (middle and high schools). They also suggest that more prompt isolation and testing could have hindered the spread, stressing how important timeliness is in this context. Despite their opposing positions, most researchers emphasize the need for the same measures: an active case finding approach with systematic and thorough testing of students and personnel, to understand and prevent the spread inside school.

Following this scientific debate, this work focuses on Italy, looking at the contagion curves and relating them to the dates schools opened in each of the 21 regions. Italy is facing a hard time being hit by a second wave of CoViD-19 cases that are bringing the healthcare system to its knees. This second wave started during the autumn, somewhere around the start of October 2020, and peaked in November when the government imposed a new form of lockdown with color-coded zones, based on risk in every region. September was a crucial period, as with the end of summer many activities were going back to normal, and the virus prevalence in the nation was quite low [20]. In the first days of that month, we began to see a slight increase in the number of new cases in most regions, probably due to the cross-regional movement for the summer holidays [21]. This prompted many to warn of the arrival of the second wave, but the number of new cases stabilized in the coming days, and the growth was considered small in any case. School reopened between the third and the fourth week of September [22,23] while people had also already started going back to their offices and activities. Another thing to notice is the referendum of September 20^th^ that in some regions corresponded with other elections for local government and senate representatives. While attendance was quite low, one could wonder the effects of such an event.

Because of the shocking scarcity of available and reliable data on SARS-CoV-2 infections inside Italian schools, we chose another perspective in order to investigate the hypothesis of an association between schools and the resurgence of the virus in Italy, by analyzing the growth rate of the total number of SARS-CoV-2 infections in all the Italian regions, before and after the school reopening.

## Methods

Given the scarce availability of data collected in schools that could better describe their role in Italy (for the reasons stated in the Introduction), we decided to work with the population-wide data at the regional level. We fit a piecewise linear regression model where the dependent variable is only the number of new daily confirmed CoViD-19 cases and the independent variable is just the number of days since September 1^st^ (until October 31^st^). The result is a model comprised of a changepoint and two segments, whose slopes represent the growth rate before and after the start of the exponential growth of the second wave. To have a measurement of the uncertainty in our estimates we decided to use a Bayesian framework for the regression as described in [24]. Two transformations were applied to the initial data. First, we used a 7-day rolling average as the raw data presents a weekly seasonality due to the way CoViD-19 tests are carried out and registered. Second, we applied a natural logarithm so that the exponential growth appears as an easily identifiable slope. Using the implementation described in [25], we estimated a piecewise linear regression model for each region. We are modeling our dependent variable ln(*y*) (the natural log of confirmed daily cases) as a Normal distribution whose mean depends on the regression coefficients *a*_*1*_ and *b*_*1*_ (the intercept and angular coefficient) before a changepoint Δ, and *a*_*2*_ and *b*_*2*_ after the changepoint, as represented in the formula below.

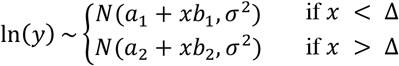

The distribution of *b*_*1*_, *b*_*2*_, *a*_*1*_, *σ* and *Δ* are estimated using Bayes theorem and Monte Carlo-Markov Chain methods. Since the two lines are joined at the changepoint, without discontinuities, the second intercept term *a*_*2*_ is not estimated as is bound to be *a*_2_ = *Δ*(*b*_1_ − *b*_2_) + *a*_1_.

As suggested in [25], the intercept and slope priors used for starting the Bayesian estimation were chosen in Gaussian families, while for the changepoint a uniform was used, precisely as reported below:

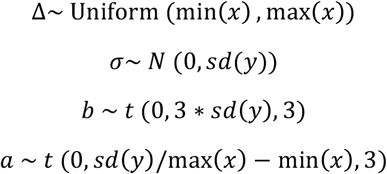

The final estimates for all the parameters (the changepoint, the two slopes, the initial intercept, and the variance) are reported in Table 1 with the 95% CI. To give an idea of the increase in slope, we computed the number of days (*DT*_*1*_ and *DT*_*2*_) necessary to observe a doubling in the number of new cases from the changepoint onwards, with both slopes (using the average angular coefficient), as shown in the following equations.

**Table 1:** Estimated parameters from the 21 Italian regions along with school opening date (**Open**), number of days between opening and changepoint dates (**D**) and the doubling time for the two slopes (**DT**_**i**_). Between brackets are the 95% CI. To be noticed is that, as to P.A. Trento, the school reopening date was that of kindergarten (3/09). Using the date of 14/09 would not have changed the main result of the correspondent analysis.

| Region | Open | D | $\Delta$ | $b_1$ | $b_2$ | $a_1$ | $\sigma$ | $DT_1$ | $DT_2$ |
| --- | --- | --- | --- | --- | --- | --- | --- | --- | --- |
| Basilicata | 24/09 | -4 | 19.25 (0.01, 51.02) | 0.02 (-0.00, 0.06) | 0.06 (0.04, 0.08) | 1.15 (0.78, 1.45) | 0.35 (0.29, 0.29) | 30.10 | 11.98 |
| Campania | 24/09 | -2 | 20.85 (18.81, 22.86) | 0.00 (-0.01, 0.01) | 0.07 (0.07, 0.08) | 5.07 (5.00, 5.15) | 0.08 (0.07, 0.07) | ∞ | 9.49 |
| Abruzzo | 24/09 | 0 | 22.54 (20.73, 24.28) | -0.01 (-0.02, 0.00) | 0.10 (0.09, 0.11) | 2.73 (2.58, 2.88) | 0.18 (0.15, 0.15) | ∞ | 6.94 |
| Sardegna | 22/09 | 0 | 21.26 (17.65, 26.73) | 0.00 (-0.01, 0.01) | 0.05 (0.04, 0.05) | 3.94 (3.85, 4.03) | 0.08 (0.07, 0.07) | ∞ | 15.16 |
| Puglia | 24/09 | 8 | 30.92 (29.63, 32.20) | 0.01 (0.01, 0.01) | 0.07 (0.07, 0.07) | 4.13 (4.08, 4.19) | 0.08 (0.07, 0.07) | 69.76 | 9.76 |
| Calabria | 24/09 | 9 | 31.67 (29.68, 33.65) | -0.01 (-0.01, 0.00) | 0.10 (0.09, 0.11) | 2.75 (2.62, 2.88) | 0.19 (0.16, 0.16) | ∞ | 6.75 |
| Valle d'Aosta | 14/09 | 13 | 26.22 (24.09, 28.29) | 0.00 (-0.02, 0.01) | 0.14 (0.13, 0.15) | 0.68 (0.49, 0.87) | 0.27 (0.22, 0.22) | ∞ | 4.95 |
| Umbria | 14/09 | 14 | 26.87 (25.72, 28.09) | 0.01 (0.00, 0.01) | 0.10 (0.10, 0.10) | 2.79 (2.72, 2.87) | 0.11 (0.09, 0.09) | 70.71 | 6.87 |
| Friuli Venezia Giulia | 16/09 | 16 | 30.91 (29.49, 32.22) | 0.01 (0.01, 0.02) | 0.09 (0.08, 0.09) | 2.96 (2.88, 3.05) | 0.13 (0.11, 0.11) | 33.96 | 6.45 |
| Piemonte | 14/09 | 16 | 28.58 (27.40, 29.73) | 0.02 (0.02, 0.02) | 0.11 (0.10, 0.11) | 3.99 (3.94, 4.05) | 0.08 (0.06, 0.06) | 61.36 | 8.94 |
| Veneto | 14/09 | 16 | 29.20 (27.09, 31.62) | 0.01 (0.01, 0.02) | 0.08 (0.07, 0.08) | 4.75 (4.68, 4.82) | 0.09 (0.07, 0.07) | 51.44 | 7.74 |
| Lombardia | 14/09 | 17 | 30.30 (29.56, 31.02) | -0.01 (-0.01, 0.00) | 0.12 (0.12, 0.13) | 5.49 (5.44, 5.55) | 0.08 (0.07, 0.07) | ∞ | 5.56 |
| Toscana | 14/09 | 17 | 29.67 (28.81, 30.51) | 0.01 (0.01, 0.01) | 0.10 (0.10, 0.10) | 4.39 (4.33, 4.45) | 0.08 (0.07, 0.07) | 70.17 | 6.92 |
| Emilia-Romagna | 14/09 | 18 | 30.65 (30.11, 31.23) | -0.01 (-0.01, -0.01) | 0.09 (0.09, 0.09) | 4.86 (4.83, 4.90) | 0.05 (0.04, 0.04) | ∞ | 7.67 |
| Marche | 14/09 | 19 | 32.35 (30.53, 34.01) | 0.02 (0.01, 0.02) | 0.10 (0.09, 0.11) | 2.73 (2.63, 2.83) | 0.15 (0.12, 0.12) | 39.67 | 6.93 |
| Liguria | 14/09 | 20 | 32.74 (30.35, 35.16) | 0.02 (0.02, 0.03) | 0.08 (0.08, 0.09) | 3.85 (3.75, 3.96) | 0.15 (0.12, 0.12) | 27.91 | 8.29 |
| Molise | 14/09 | 22 | 35.16 (33.16, 37.35) | 0.02 (0.01, 0.03) | 0.12 (0.11, 0.14) | 0.72 (0.55, 0.91) | 0.29 (0.24, 0.24) | 19.63 | 9.67 |
| Sicilia | 14/09 | 22 | 34.52 (32.17, 36.81) | 0.04 (0.03, 0.04) | 0.07 (0.07, 0.08) | 3.81 (3.75, 3.87) | 0.09 (0.08, 0.08) | 36.39 | 5.71 |
| Lazio | 14/09 | 23 | 35.85 (34.69, 37.01) | 0.02 (0.01, 0.02) | 0.09 (0.09, 0.10) | 4.89 (4.85, 4.94) | 0.07 (0.05, 0.05) | 41.98 | 7.60 |
| P.A. Bolzano | 7/09 | 28 | 34.39 (29.46, 41.99) | 0.04 (0.03, 0.05) | 0.10 (0.07, 0.12) | 1.97 (1.83, 2.12) | 0.22 (0.17, 0.17) | 17.79 | 6.87 |
| P.A. Trento | 3/09 | 43 | 44.99 (42.97, 47.17) | 0.02 (0.02, 0.03) | 0.12 (0.09, 0.14) | 2.68 (2.55, 2.81) | 0.24 (0.20, 0.20) | 32.14 | 5.90 |

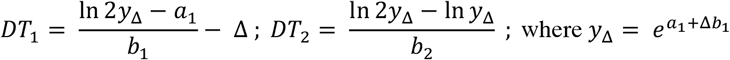

All the data used are available from the Italian government site: https://github.com/pcm-dpc/COVID-19. The results of all the computations are fully reproducible by using the method stated above.

### Patient and Public Involvement

Patients and/or the public were not involved in the design, or conduct, or reporting, or dissemination plans of this research.

## Results

Table 1 shows all the estimated quantities for each region. Similarly, Figure 1 shows all the curves and regression lines. Out of the 21 Italian regions, 15 (71%) of them have a changepoint within 28 days from the date when the school opened.In particular, the average number of days between the opening and the changepoint is 16.66 days (CI 95% 14.47 to 18.73). We believe this number is plausible in a scenario where one has to be exposed to the virus and then manifest symptoms in order to be tested, also considering that often children and adolescents are asymptomatic and that they only function as a vector to other people around them.

**Figure 1:**
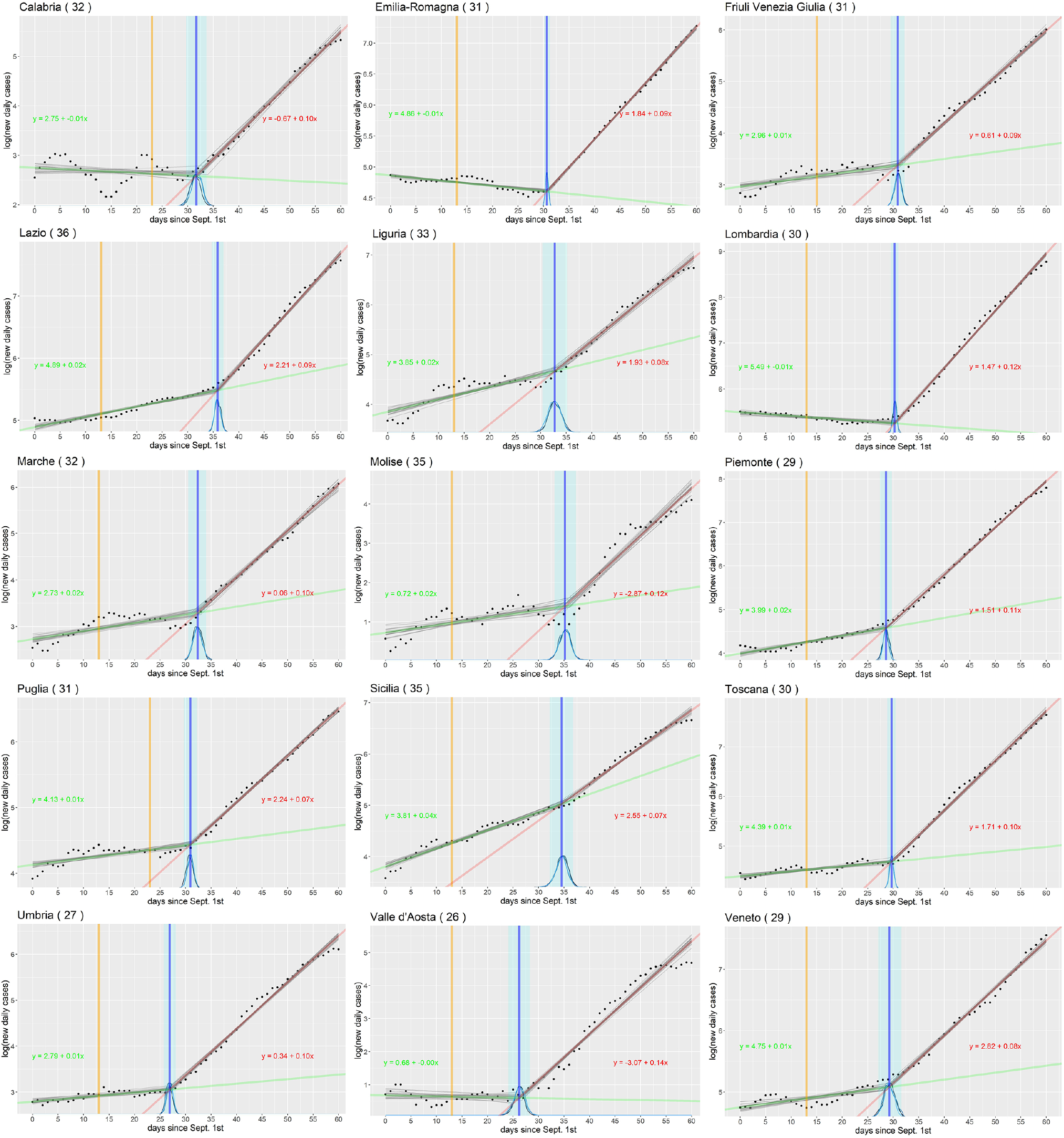
Plots for the regions whose changepoint (blue with 95% CI area) is within 28 days since school openings (yellow). Black dots are the natural log of daily confirmed <u>CoViD</u>-19 cases from 09/01/2020 to 31/10/2020. In Green and Red the regression lines using the mean parameters, before and after the changepoint.

Looking at the estimated slopes of Figure 1, we can have an idea of the strength of the increase. While one could comment on the difference in the angular coefficients, converting it into an angle, the number does not convey the idea very effectively. Translating it into the number of days required for a doubling in the growth rate is more interpretable. Of the 15 regions mentioned above, 4 had a slope that is null or slightly negative before the change, making this time infinite, but the fact that the trend was inverted to something like 7 days is significant enough by itself. The remaining 11 regions went from an average of 47.50 (CI 95% 37.18 to 57.61) days at the rate before the changepoint to an average of 7.72 (CI 95% 7.00 to 8.48) at the rate after the changepoint. Table 1 and Figure 1 illustrate these results.

Of the 6 regions that break the pattern, two of them, P.A. Trento and P.A. Bolzano (often considered one region, called Trentino-Alto Adige) presented a changepoint more than four weeks after the school opening; they are indicated in blue in Table 1. On the other hand, the remaining four, in yellow in Table 1, begin their exponential rise before or in correspondence of the school opening. Both are displayed in Figure 2.

**Figure 2:**
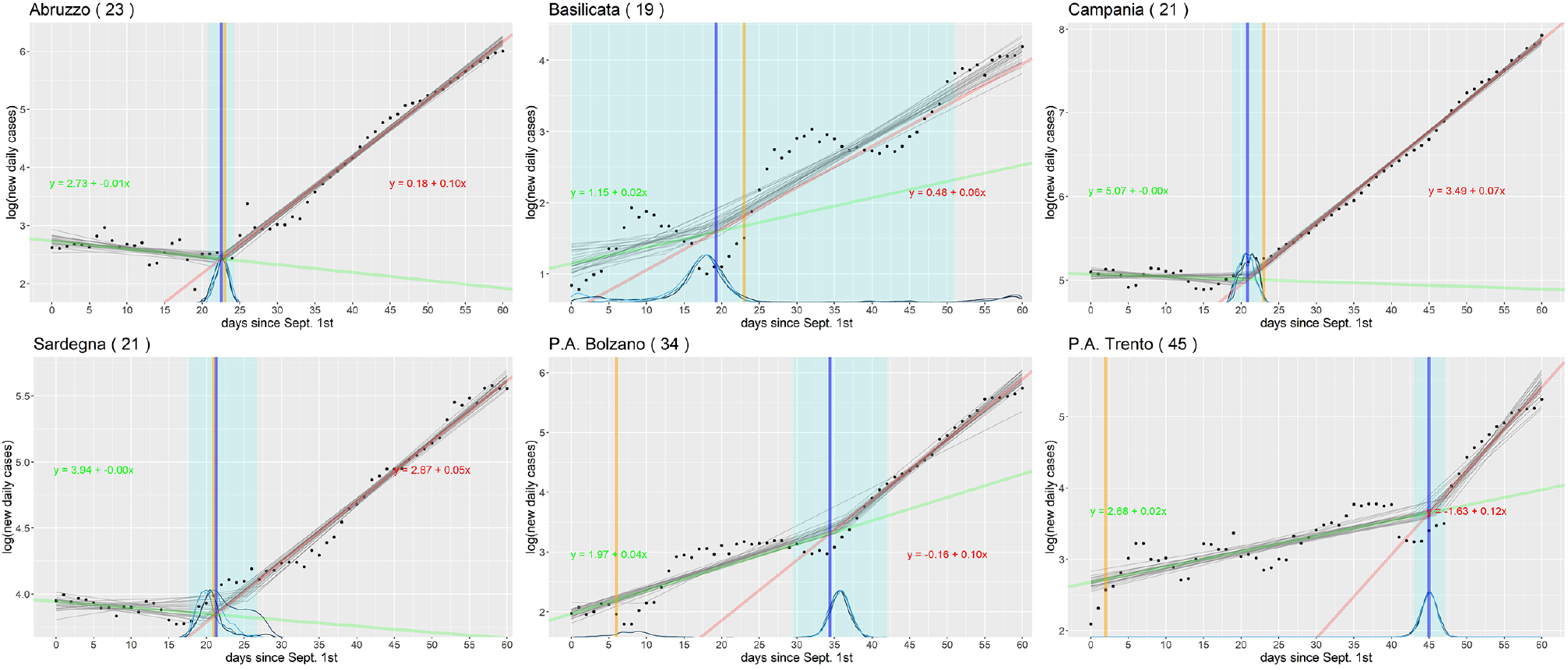
The same plots of Figure 1 but for the regions that do not exhibit the pattern. First four anticipate the start of the exponential growth before school open, the last two more take than 28 days after that.

All the models converge quite nicely, with narrow 95% CI, except for Basilicata, where the changepoint estimation is extremely wide, probably due to the high variability in the reported numbers.

## Discussion

We can surely conclude from the results that something has triggered the second wave in September in Italy. Certainly, multiple confounding factors played a role in the acceleration of the SARS-CoV-2 infection, but our opinion is that schools are surely one of those, and the magnitude of their effects should be investigated more thoroughly.

In the short period that precedes the second wave there were not many events that interested as many people as schools. The referendum and elections on September 20th almost coincide with school opening and did not have a large participation. Workers going back to their offices could also have played a major role, however by looking at the nation-wide mobility report by Google, we can see that in September the number of people moving to their workplace increased steadily from −30% to −20% with respect to the reference level before the pandemic. If we consider that in Italy there are approximately 25 million workers, an increase of 10% would translate into 2.5 million more people circulating. If we compare this number with that of students and school personnel which is equal to approximately 11 million, we can conclude that perhaps, even when considering that both categories use public transportation heavily, schools could be more influential in spreading the virus.

The regions that do not follow the pattern may tell us something more. Trento and Bolzano could be outliers, as they are often considered a single region and likely share characteristics that could set them apart from other regions. In the others, the change is before or coincidental with school opening, so any factor that could have ignited the second wave is to be researched outside of the school activity and all the other connected activities. The effect of schools, if any, would be absorbed in the inflation already in act. being all four maritime regions, tourism could perhaps be one of the major causes.

Regarding the regions where the number of cases has high variability, the reason is most likely that the number of tests done each day varies as much. We could have normalized the cases with the number of tests, but this datum is often unreliable and leads to unrealistic normalized values, so we decide to avoid this.

As a final note, any research hypothesis concerning SARS-CoV-2 infections in schools has a hard time being verified in Italy as no region has so far seriously investigated the dynamics of the spread of the virus inside schools by using a controlled and systematic active case finding methodology.

## Conclusion

We have applied a piecewise regression to the number of new daily CoViD-19 cases since September 1^st^, 2020 in order to highlight the start of the second wave and relate it to the reopening of schools in Italy. The numbers emerged from this study are not enough to rule out schools, as some suggest, and neither they indicate a direct link, as others are sure of. Nonetheless they show that the exponential growth is compatible, most of the time, with schools opening two to three weeks prior. We believe that a serious controlled study is necessary, to give a definitive answer to this question, with a comparison between different age brackets and schools, and an effort to track the virus inside and outside the school’s walls, using an active approach to case finding. The difference between regions, especially those that break the pattern, should be investigated as it could provide clues on how the dynamics of the spread change according to context.

## Data Availability

Data are available in a public, open access repository (https://github.com/pcm-dpc/COVID-19). 
All data and code are available upon request to the corresponding author

